# Predictors and rates of PTSD, depression and anxiety in UK frontline health and social care workers during COVID-19

**DOI:** 10.1101/2020.10.21.20216804

**Authors:** Talya Greene, Jasmine Harju-Seppänen, Mariam Adeniji, Charlotte Steel, Nick Grey, Chris R. Brewin, Michael A. Bloomfield, Jo Billings

**Author notes:** **Corresponding author is Talya Greene**.

## Abstract

**Background:** Studies have shown that working in frontline healthcare roles during epidemics and pandemics was associated with PTSD, depression, anxiety, and other mental health disorders.

**Objectives:** The objectives of this study were to identify demographic, work-related and other predictors for clinically significant PTSD, depression, and anxiety during the COVID-19 pandemic in UK frontline health and social care workers (HSCWs), and to compare rates of distress across different groups of HCSWs working in different roles and settings.

**Methods:** A convenience sample (*n=1194*) of frontline UK HCSWs completed an online survey during the first wave of the pandemic (27 May – 23 July 2020). Participants worked in UK hospitals, nursing or care homes and other community settings. PTSD was assessed using the International Trauma Questionnaire (ITQ); Depression was assessed using the Patient Health Questionnaire-9 (PHQ-9); Anxiety was assessed using the Generalized Anxiety Disorder Scale (GAD-7).

**Results:** Nearly 58% of respondents met the threshold for clinically significant PTSD, anxiety or depression, and symptom levels were high across occupational groups and settings. Logistic regression analyses found that participants who were concerned about infecting others, who felt they could not talk with their managers, who reported feeling stigmatised and who had not had reliable access to personal protective equipment (PPE) were more likely to meet criteria for a clinically significant mental disorder. Being redeployed during the pandemic, and having had COVID were associated with higher odds for PTSD. Higher household income was associated with reduced odds for a mental disorder.

**Conclusions:** This study identified predictors of clinically significant distress during COVID-19 and highlights the need for reliable access to PPE and further investigation of barriers to communication between managers and staff.

## Introduction

Since the COVID-19 global pandemic began, frontline health and social care workers (HSCWs) have been repeatedly identified as being at high risk for severe psychological distress (Billings, Greene, et al., 2020; Gersons, Smid, Smit, Kazlauskas, & McFarlane, 2020; Greenberg, Docherty, Gnanapragasam, & Wessely, 2020; Javakhishvili et al., 2020; Shanafelt, Ripp, & Trockel, 2020), and emerging research seems to support this (Billings, Ching, Gkofa, Greene, & Bloomfield, 2020; Braquehais et al., 2020; Lai et al., 2020; Pappa et al., 2020). The COVID-19 situation is dynamic within, and variable across, countries with a range of health care systems. It is important to examine mental distress among HSCWs in different countries at different phases of the pandemic to guide national responses as well as learn from the international context. Evidence is required not only to evaluate how widespread mental distress is among frontline HSCWs but, crucially, to identify risk factors. This will help identify which frontline health and social care workers are at highest risk, inform evidence-based primary prevention strategies during anticipated subsequent peaks associated with COVID-19 and guide secondary prevention treatment strategies to minimise distress, as well as increase understanding of occupational stressors for HSCWs in general.

During a pandemic, frontline HSCWs are at increased risk of infection while also dealing with working conditions that are more difficult and demanding than usual (Shanafelt et al., 2020). Studies have shown that working in healthcare roles during epidemics and pandemics such as SARS, MERS, and Ebola disease, was associated with PTSD, depression, anxiety, and other mental health disorders (Allan et al., 2020; Brooks, Dunn, Amlôt, Rubin, & Greenberg, 2018; de Pablo et al., 2020; Preti et al., 2020). Research has also highlighted potential risk factors, including being tasked to directly work with patients suspected of having the virus (Kisely et al., 2020; Maunder et al., 2003; McAlonan et al., 2007; Styra et al., 2008) working as a nurse (Billings, Ching, et al., 2020; de Pablo et al., 2020; Preti et al., 2020), being a parent of dependent children (Kisely et al., 2020) and being a woman (Serrano-Ripoll et al., 2020). HCSWs may have concerns both about being infected, and about passing the disease onto others (Bai et al., 2004; Billings, Ching, et al., 2020), and may feel stigmatised as a result of their role (Bai et al., 2004; Brooks et al., 2018; de Pablo et al., 2020).

Emerging evidence from research on the COVID-19 pandemic indicates high rates of mental disorders among HSCWs in many countries, including China, USA, India, and Italy (Braquehais et al., 2020; Carmassi et al., 2020; Chew et al., 2020; Krishnamoorthy, Nagarajan, Saya, & Menon, 2020; Pappa et al., 2020; Shechter et al., 2020). Studies have identified potential risk factors (Braquehais et al., 2020; Lai et al., 2020), especially concerns about personal safety given the elevated rates of morbidity and mortality among healthcare workers which may have been exacerbated where there was inadequate access to appropriate personal protective equipment (PPE) (Braquehais et al., 2020; Urooj, Ansari, Siraj, Khan, & Tariq, 2020; Wang, Zhou, & Liu, 2020), and among staff directly working with patients with confirmed or suspected COVID-19 (Kang et al., 2020). Additionally, being a woman (Lai et al., 2020) and working as a nurse (Luceño-Moreno, Talavera-Velasco, García-Albuerne, & Martín-García, 2020) were both associated with higher mental distress. Staff may feel they have provided suboptimal treatment due to insufficient staff and resources, and working with social restrictions, which may cause distress sometimes referred to as moral injury (Williamson, Murphy, & Greenberg, 2020). In some locations and settings (e.g., Northern Italy) there was a need to ration treatments which has been especially challenging for healthcare workers, compounding an already difficult situation (Cavallo, Donoho, & Forman, 2020).

In England, it has been estimated that one in ten COVID-19 cases around the peak of the first wave occurred in frontline HSCWs (Torjesen, 2020). A survey of UK HSCWs published in April 2020 found that half of those surveyed reported that their mental health had deteriorated during the pandemic (Thomas, Quilter-Pinner, & Research, 2020). In a survey of UK doctors in May 2020, 45% reported experiencing depression, anxiety, stress, burnout or other mental health problems during the pandemic (BMA, 2020), and a large survey of UK nurses published in August 2020 found that 76% reported an increase in their stress since the outbreak of COVID-19, and over half were concerned about their mental health (Royal College of Nursing, 2020). It is clear that working in health and social care during COVID-19 in the UK has been challenging. To date, however, studies have not been published assessing rates of PTSD, depression, and anxiety among different UK HSCW groups during COVID-19, nor have any studies been published examining risk factors for these mental health disorders during the pandemic in the UK.

The existing studies, both those examining COVID-19 related distress, and those from previous pandemics, have focused predominantly on medical staff (nurses and doctors). Some have also included allied health professionals but have rarely included clinical support staff or auxiliary frontline healthcare workers such as receptionists, porters and cleaners. Importantly, the psychological impact of pandemics on social care workers has been largely neglected, yet care homes have been particularly badly affected by COVID-19 (Gordon et al., 2020). Studies based on wider samples are urgently needed.

The Frontline-COVID study is an online survey for which baseline data were collected during the first wave of the COVID-19 pandemic. The study aimed to identify demographic, work-related and other predictors for psychological distress (clinically significant levels of PTSD/CPTSD, depression, and/or anxiety) during the pandemic in UK frontline HSCWs, and to compare rates of PTSD, depression and anxiety across different groups of HCSWs working in a wide variety of roles, including clinical, non-medical, allied healthcare and auxiliary roles.

## Methods

### Participants and procedure

Frontline health and social care workers across the UK were invited to participate in the study via a social media campaign (Facebook adverts, Twitter and Facebook posts, and emails to wellbeing leads at a number of UK hospitals, with a request to circulate to staff). The questionnaire was administered using online survey methods, via the Qualtrics data collection platform. Data were collected between 27 May 2020-23 July 2020. This represents the post-peak phase of the initial COVID-19 wave in the UK; during this period, deaths related to COVID in the UK rose from 37,430 to 41,160, while reported weekly deaths fell from 2000 (29 May 2020) to 231 (24 July 2020) (Public Health England, 2020). Participants gave informed consent online before proceeding to the questionnaire. Ethical approval for the Frontline-COVID study was granted by the UCL Ethics Committee.

In total, 2447 individuals opened the link to read the participant information sheet, 1311 consented to participate, and 1205 provided data. Participants who indicated that they did not work in healthcare *(n= 5*) were excluded. In cases where participants completed the questionnaire on more than one occasion, the first response was used and the second was excluded from analysis (*n= 6*). This resulted in a sample of 1194 individuals.

### Measures

The study survey included background questions regarding participants’ gender, age, income, ethnicity, whether they were in a relationship, whether they were caring for children at home, and UK region of work. Participants were asked to indicate their job role which was then operationalised into the following categories: Nurse or midwife; carer (mostly working in care home or community settings); clinical support staff (including healthcare assistants); doctor; non-clinical staff working in health and social care settings (including cleaners, porters, administrators, maintenance, security roles); allied healthcare professionals (including physiotherapists, occupational therapists, paramedics, and other allied roles as defined by the NHS); and any other roles. Participants also reported their work setting, which was operationalised as follows: Hospital, nursing or care home, and any other community setting. A series of questions assessed: access to PPE (yes vs no or sometimes); whether they were redeployed during COVID-19 (currently or previously redeployed vs no); whether they had been infected with COVID-19 (confirmed or suspected vs no); using alcohol, cigarettes or other substances more than usual to cope (yes vs no); and whether they could tell their manager or team leader if they were not coping (yes vs no). Single Likert scales ranging from 0 (“not at all”) to 4 (“extremely”) assessed: whether participants worried they would be rejected or stigmatized for being an NHS worker; whether they were worried about being infected with COVID-19; and whether they were worried about infecting others with COVID-19.

#### PTSD symptoms

were assessed using the International Trauma Questionnaire (ITQ; Cloitre et al., 2018). This is a self-report questionnaire, based on the ICD-11 criteria for PTSD and Complex PTSD (CPTSD), which has demonstrated reliability and validity. As all those meeting criteria for CPTSD also need to meet criteria for PTSD, we only examined the PTSD threshold in order to identify those with clinical levels of distress. For the identification of participants meeting PTSD thresholds, individuals report how often they have experienced six core symptoms of PTSD (two from each of three subscales) in the last month and three functional impairment items related to these subscales, on a 5-point Likert scale ranging from 0 (“not at all”) to 4 (“extremely”). The diagnostic threshold for PTSD is met if at least one of two symptoms from each PTSD symptom subscale are endorsed (scored as 2:: 2) and there is endorsement of at least one of the functional impairment items. In total, 1110 individuals completed the first nine items of the ITQ. Cronbach’s alpha in the current study was 0.88.

#### Depression symptoms

were assessed using the Patient Health Questionnaire-9 (Kroenke, Spitzer, & Williams, 2001) (PHQ-9). This is a widely used 9-item self-report questionnaire corresponding to the DSM-5 criteria for depression. Previous studies indicate that the PHQ-9 has high internal consistency and test-retest reliability (Kroenke et al., 2001). In the current study, participants reported how often symptoms occurred during the previous fortnight on a four-point Likert scale ranging from 0 (‘not at all’) to 3 (‘very much’). Total scores are the sum of all item scores (0 - 27); a score of 10 or higher typically indicates moderate depression. Altogether, 1017 individuals completed the PHQ-9. Cronbach’s alpha in the current study was 0.90.

#### Anxiety symptoms

were assessed using the Generalised Anxiety Disorder Scale-7 (Spitzer, Kroenke, Williams, & Löwe, 2006) (GAD-7). This is a 7-item self-report questionnaire developed to indicate severity of anxiety symptoms and has demonstrated high internal consistency and test-retest reliability. Participants report how much they have been bothered by each symptom over the past two weeks on a 4-point Likert Scale ranging from 1 (“not at all”) to 3 (“more than half the days”). Total scores are the sum of all item scores (0 - 21). In UK clinical settings, a score of 8 is used for clinically significant anxiety. Altogether, 994 individuals completed the GAD-7. Cronbach’s alpha for the current study was 0.93.

### Data analysis

Four separate logistic regressions were performed. The regressions examined the predictors for depression, anxiety and PTSD/CPTSD separately, and also investigated the predictors of meeting the thresholds for at least one of the three conditions. Participants were excluded if they had missing data on any of the predictor variables. Additionally, participants were excluded if they had not completed the relevant outcome measure. This resulted in the following sample sizes: 851 for the analysis investigating if participants met the threshold for at least one of the three disorders, 943 for PTSD, 876 for depression, and 854 for anxiety. Analyses were conducted in R (version 3.6.2) using the *glm* package. The p-value threshold was set as 0.05.

## Results

The mean age of the participants was 41.5 years (range = 18.5 – 86.5; SD =11.8). Overall, the majority of the sample were female (92.4%), white (90.8%) and married or living with a partner (63%). Of the participants, 75.6% reported that they had worked directly to treat, support or care for patients with COVID, 17.7% reported having had confirmed COVID, and a further 12.9% reported having had suspected COVID. For more participant details, see Table 1. For details of numbers of individuals in each role by setting, please see supplementary material.

**Table 1.**
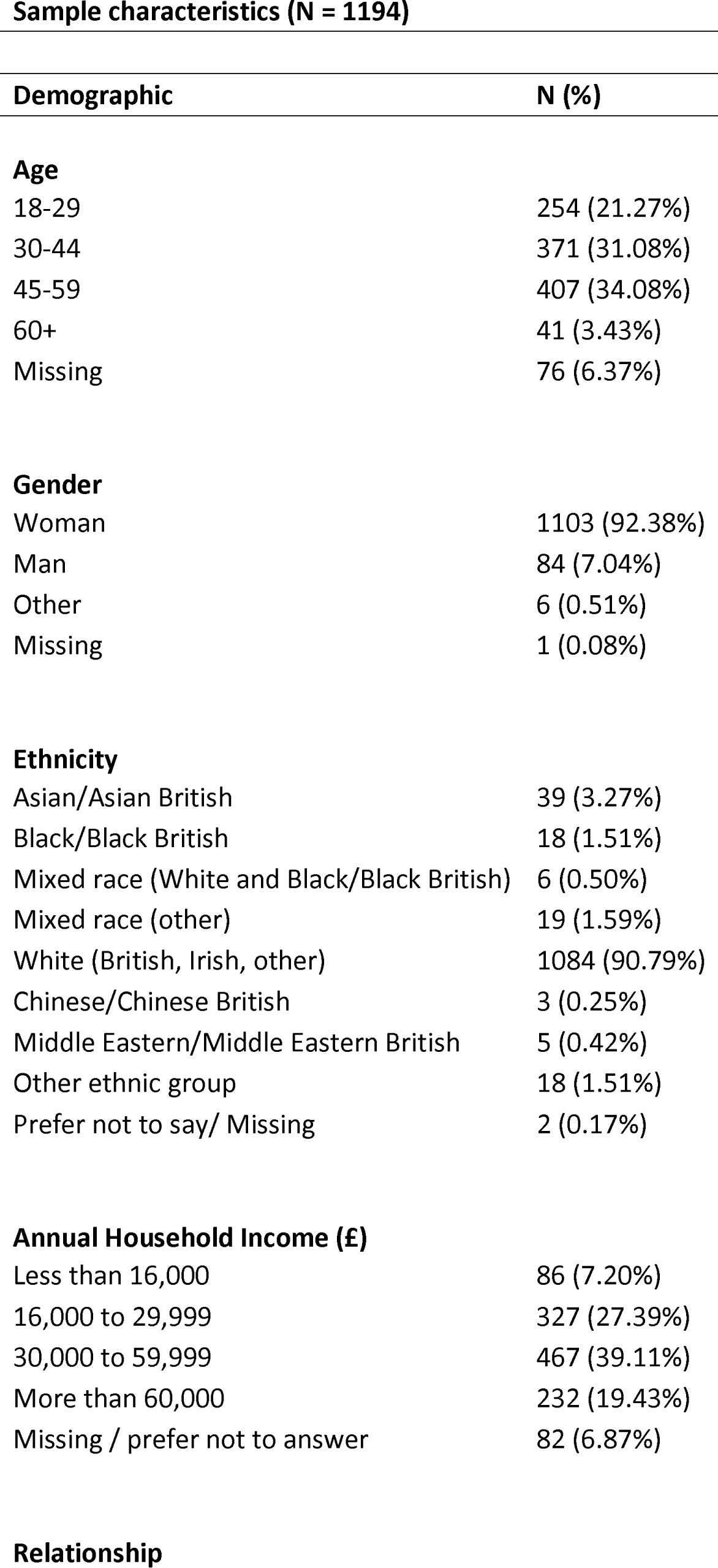
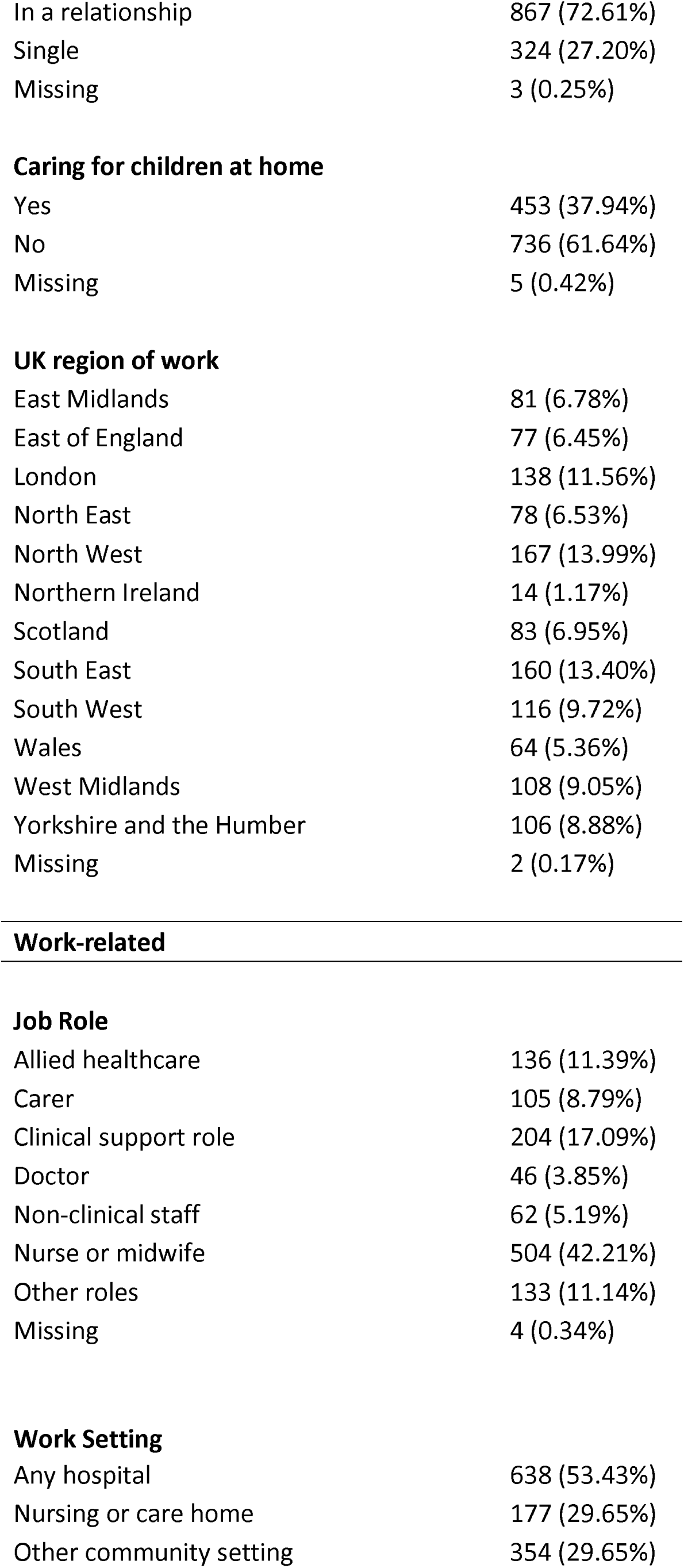
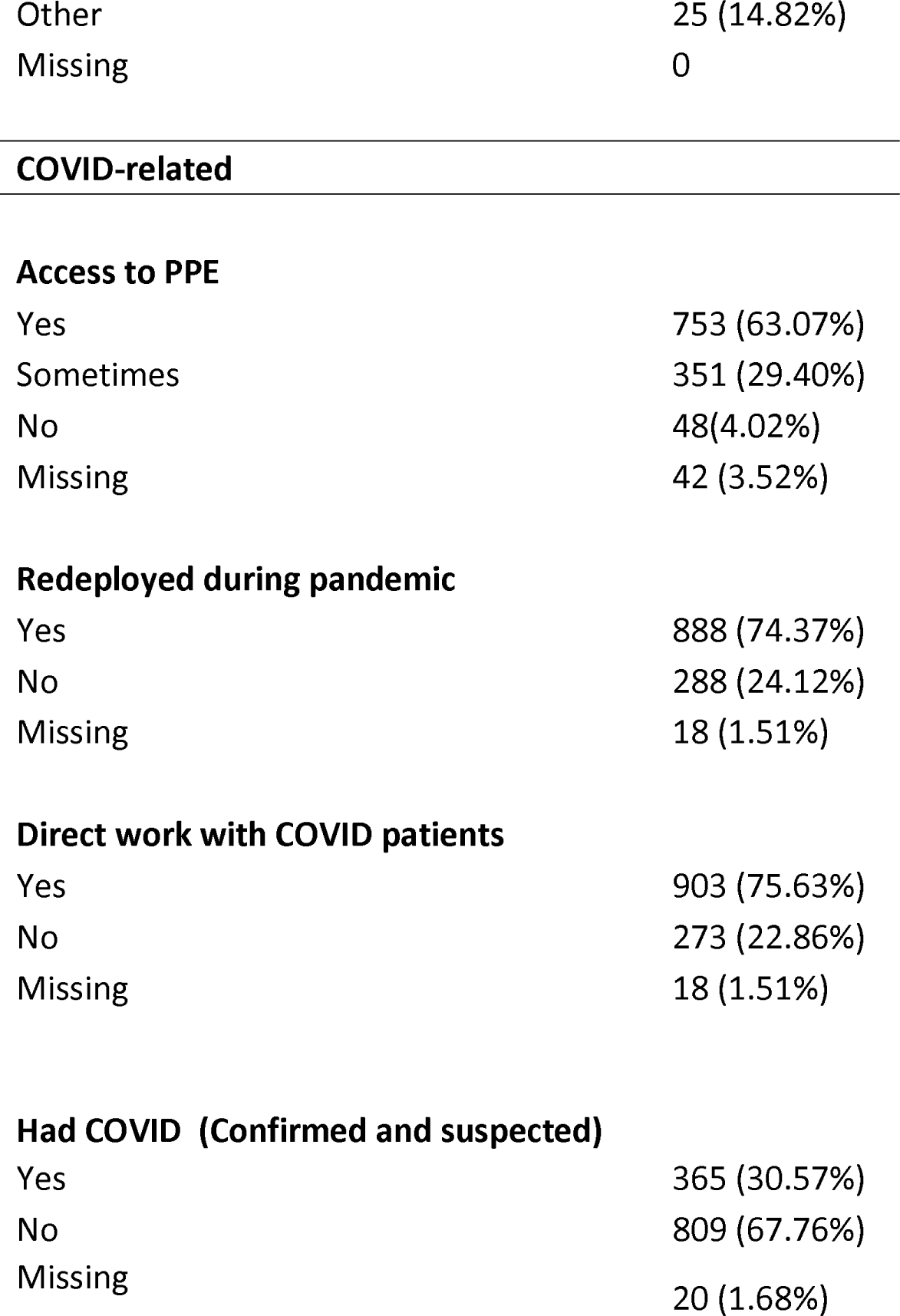
Characteristics of the study participants.

**Sample characteristics (N = 1194)**
| **Demographic** | **N (%)** |
| --- | --- |
| **Age** |  |
| 18-29 | 254 (21.27%) |
| 30-44 | 371 (31.08%) |
| 45-59 | 407 (34.08%) |
| 60+ | 41 (3.43%) |
| Missing | 76 (6.37%) |
| **Gender** |  |
| Woman | 1103 (92.38%) |
| Man | 84 (7.04%) |
| Other | 6 (0.51%) |
| Missing | 1 (0.08%) |
| **Ethnicity** |  |
| Asian/Asian British | 39 (3.27%) |
| Black/Black British | 18 (1.51%) |
| Mixed race (White and Black/Black British) | 6 (0.50%) |
| Mixed race (other) | 19 (1.59%) |
| White (British, Irish, other) | 1084 (90.79%) |
| Chinese/Chinese British | 3 (0.25%) |
| Middle Eastern/Middle Eastern British | 5 (0.42%) |
| Other ethnic group | 18 (1.51%) |
| Prefer not to say/ Missing | 2 (0.17%) |
| **Annual Household Income (£)** |  |
| Less than 16,000 | 86 (7.20%) |
| 16,000 to 29,999 | 327 (27.39%) |
| 30,000 to 59,999 | 467 (39.11%) |
| More than 60,000 | 232 (19.43%) |
| Missing / prefer not to answer | 82 (6.87%) |
| **Relationship** |  |
| In a relationship | 867 (72.61%) |
| Single | 324 (27.20%) |
| Missing | 3 (0.25%) |
| **Caring for children at home** |  |
| Yes | 453 (37.94%) |
| No | 736 (61.64%) |
| Missing | 5 (0.42%) |
| **UK region of work** |  |
| East Midlands | 81 (6.78%) |
| East of England | 77 (6.45%) |
| London | 138 (11.56%) |
| North East | 78 (6.53%) |
| North West | 167 (13.99%) |
| Northern Ireland | 14 (1.17%) |
| Scotland | 83 (6.95%) |
| South East | 160 (13.40%) |
| South West | 116 (9.72%) |
| Wales | 64 (5.36%) |
| West Midlands | 108 (9.05%) |
| Yorkshire and the Humber | 106 (8.88%) |
| Missing | 2 (0.17%) |
| **Job Role** |  |
| Allied healthcare | 136 (11.39%) |
| Carer | 105 (8.79%) |
| Clinical support role | 204 (17.09%) |
| Doctor | 46 (3.85%) |
| Non-clinical staff | 62 (5.19%) |
| Nurse or midwife | 504 (42.21%) |
| Other roles | 133 (11.14%) |
| Missing | 4 (0.34%) |
| **Work Setting** |  |
| Any hospital | 638 (53.43%) |
| Nursing or care home | 177 (29.65%) |
| Other community setting | 354 (29.65%) |
| Other | 25 (14.82%) |
| Missing | 0 |
| **Access to PPE** |  |
| Yes | 753 (63.07%) |
| Sometimes | 351 (29.40%) |
| No | 48(4.02%) |
| Missing | 42 (3.52%) |
| **Redeployed during pandemic** |  |
| Yes | 888 (74.37%) |
| No | 288 (24.12%) |
| Missing | 18 (1.51%) |
| **Direct work with COVID patients** |  |
| Yes | 903 (75.63%) |
| No | 273 (22.86%) |
| Missing | 18 (1.51%) |
| **Had COVID (Confirmed and suspected)** |  |
| Yes | 365 (30.57%) |
| No | 809 (67.76%) |
| Missing | 20 (1.68%) |

Of the study participants, 391 (32.8%) reported that they were using alcohol, cigarettes, and other substances more than usual, and 360 (30.2%) reported that they could not tell their manager or team leader how they were feeling. There were 668 participants (56%) who reported being moderately to extremely worried about catching COVID, 927 (77.6%) were moderately to extremely worried about infecting others, and 435 (36.5%) reported feeling moderately to extremely stigmatised.

### Rates of clinically significant distress

Rates of clinically significant distress for PTSD, depression and anxiety were assessed (see Table 2). Notably, 57.9% of participants met criteria for clinically significant levels of distress.

**Table 2.** Rates for clinically significant distress.

|  |  |
| --- | --- |
| **PTSD N = 1110 for mean score, N = 1095 for diagnosis** |  |
| Mean (SD) | 7.96 (5.81) |
| PTSD present (N/%) | 246 (22.47%) |
| **Depression N = 1017** |  |
| Mean (SD) | 9.97 (6.57) |
| Depression present (N/%) | 477 (46.90%) |
| **Anxiety N = 994** |  |
| Mean (SD) | 8.49 (6.14) |
| Anxiety present (N/%) | 470(47.28 %) |
| **PTSD, Depression and/or Anxiety N = 988** |  |
| Anxiety, Depression and/or PTSD (N/%) | 572 (57.89%) |

#### Predictors

Due to the group sizes on some of the predictor variables, some categories were merged: doctors and non-clinical staff were merged with the ‘other’ category for the roles variable; ethnic background was dichotomized such that participants identifying as Black, Asian or other ethnic minorities were merged into one category, and participants identifying as White were placed in the second category.

Figure 1 shows the level of PTSD, depression and anxiety symptoms, by professional role or occupational group. ANOVAs showed that the only significant group differences were between allied healthcare professionals and clinical support staff, with clinical support staff reporting more symptoms across all three disorders. We also compared PTSD, depression and anxiety symptoms by setting, and found that the only significant difference was that participants working in nursing or care home settings had higher levels of PTSD symptoms compared with other community settings. For full analyses and for zero-order associations of other study variables please see the supplementary material.

**Figure 1:**
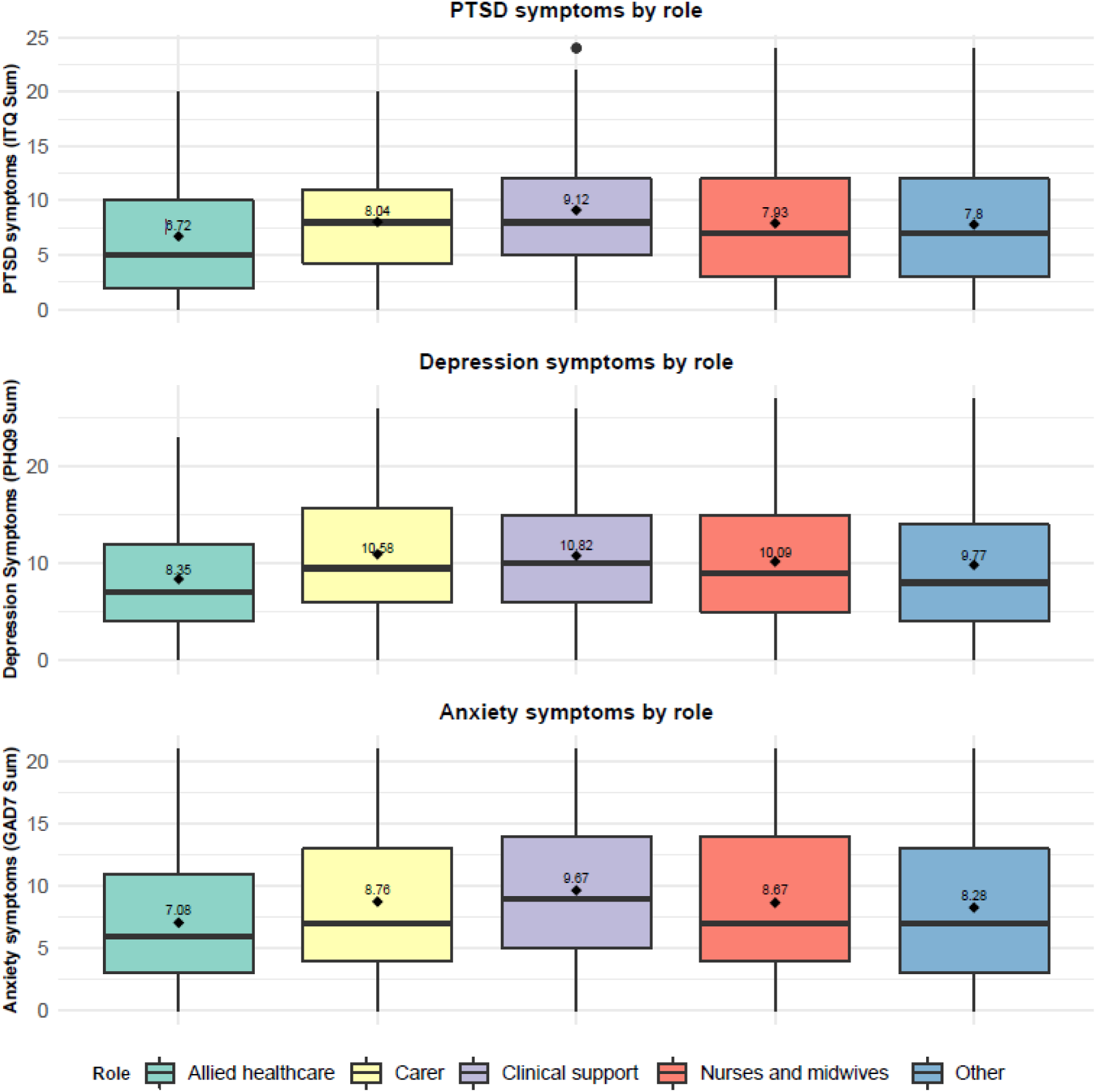
Clinically significant symptoms by role. **Figure note: The mean value is denoted by the diamond point on the box plot**

We investigated the predictors of meeting thresholds for at least one of PTSD, depression and anxiety, and also for meeting diagnostic thresholds for each disorder separately, using logistic regressions (Table 3).

**Table 3.**
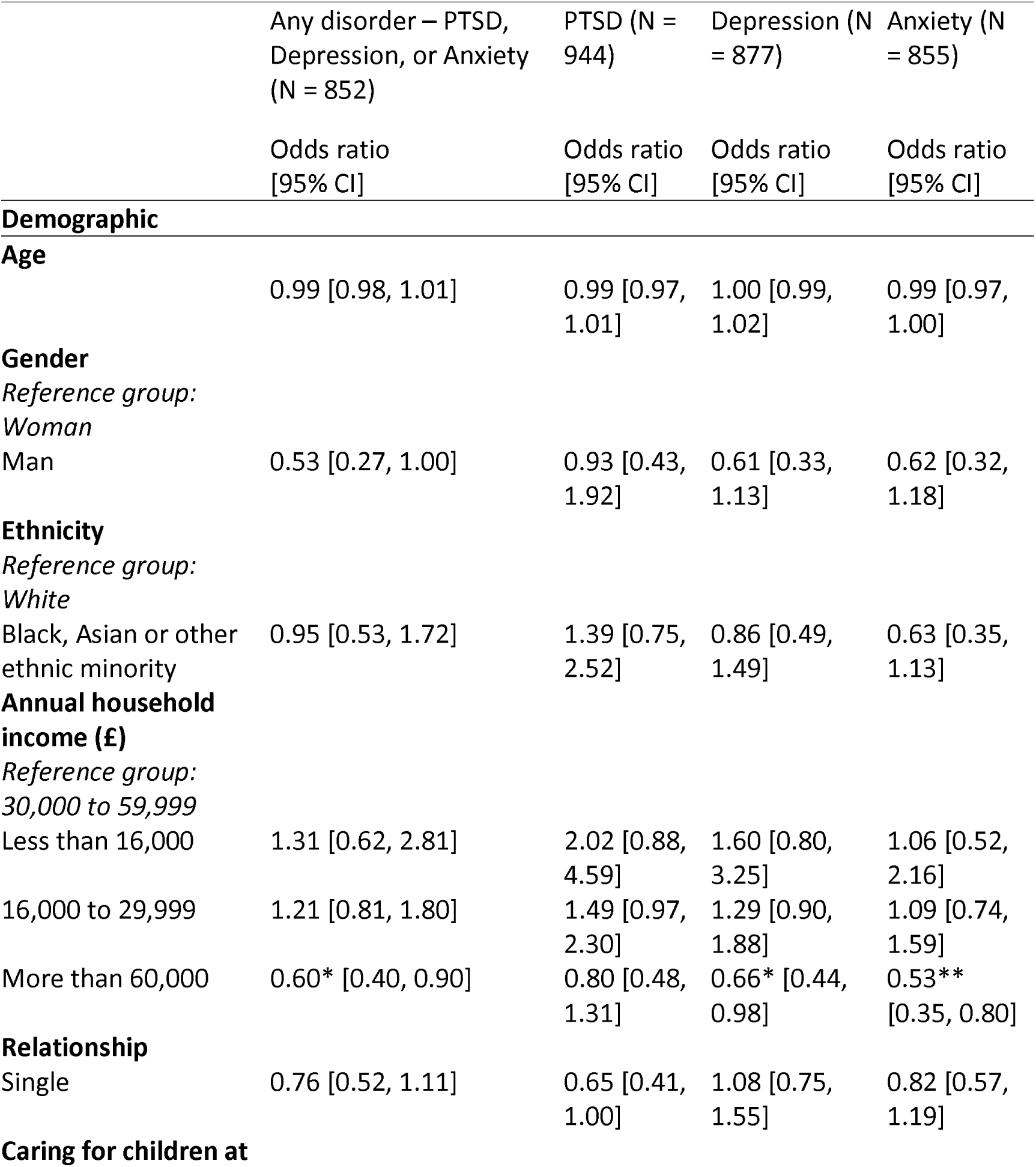
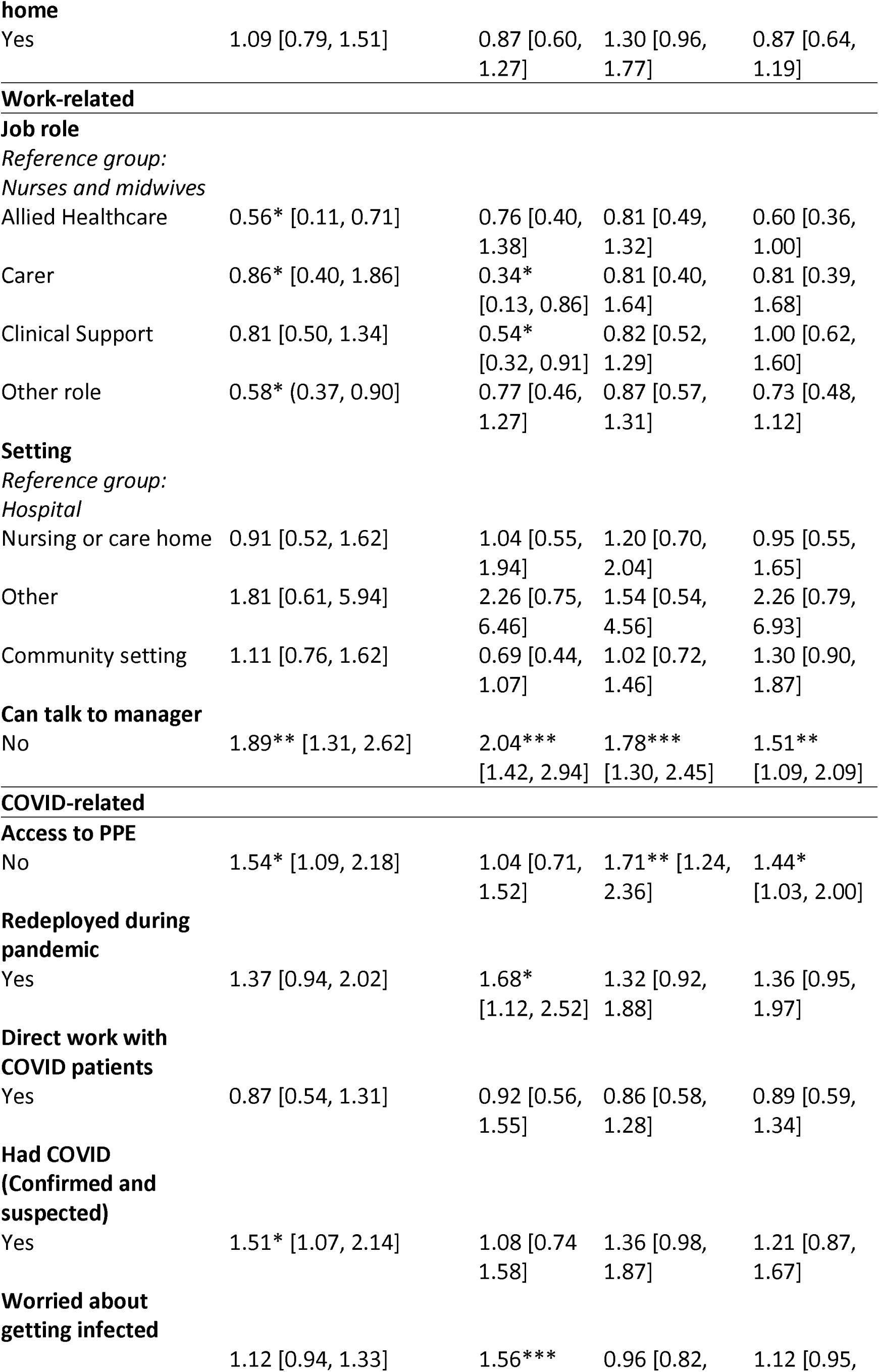
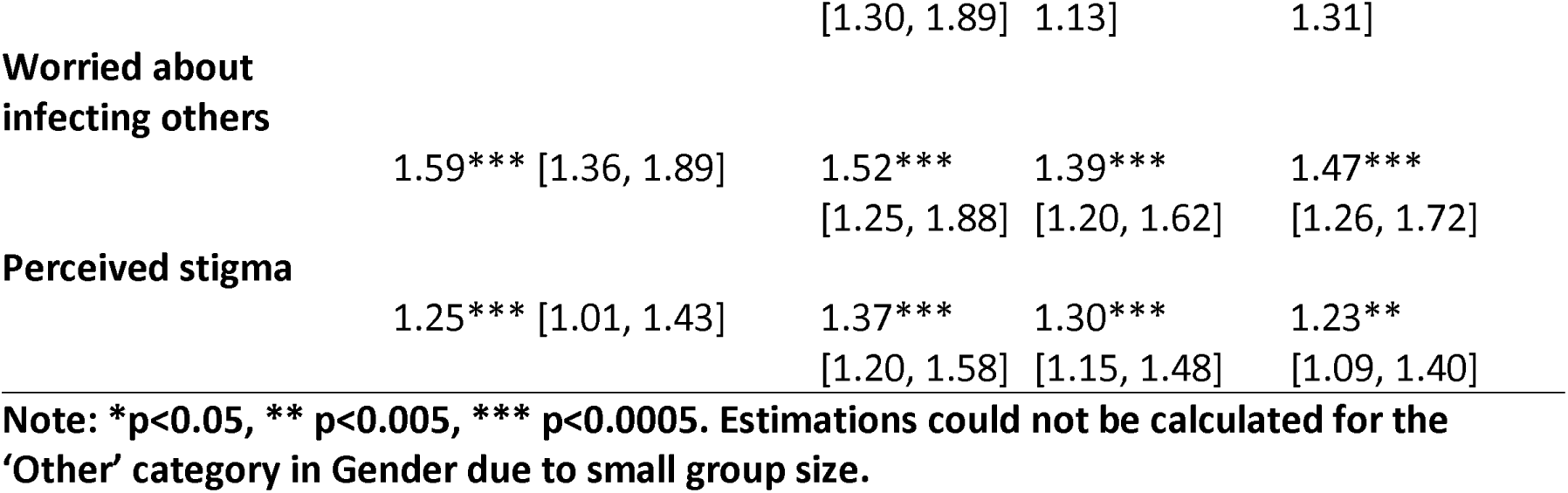
Logistic regressions for clinically significant PTSD, depression and anxiety.

Three variables significantly predicted distress across all four models: not feeling able to tell a manager they were not coping; being worried about infecting others; and perceived stigma. Participants who reported not having had reliable access to PPE had higher odds of meeting criteria for both depression and anxiety, as well as any clinically significant disorder. Having been redeployed during COVID-19 was associated with higher odds for PTSD. Nurses and midwifes were significantly more likely to meet criteria for PTSD compared with carers and clinical support staff, and they were also more likely to meet criteria for any clinically significant disorder vs allied healthcare professionals, carers, and the heterogeneous ‘other roles’ group. Participants who were worried about being infected had higher odds of meeting criteria for PTSD. Having had COVID (confirmed or suspected) was also associated with increased odds of meeting criteria for any clinically significant disorder. The group with the highest income were least likely to meet criteria for PTSD and anxiety, and also had lower odds for any clinically significant disorder.

## Discussion

This study examined rates and predictors of clinically significant PTSD, depression and anxiety in frontline health and social care workers across the UK during the first wave of the COVID-19 pandemic. Our findings indicate that clinically significant distress was common, with over 57% of respondents meeting the threshold for PTSD, anxiety and/or depression. Nearly a third of respondents reported using alcohol, cigarettes or other substances more than usual to cope. Participants who were concerned about infecting others, who felt they could not talk with their managers, who reported feeling stigmatised due to their role, who had not had reliable access to PPE, and who had caught COVID were more likely to have a clinically significant mental disorder. Being redeployed during the pandemic and being worried about catching COVID were associated with a higher likelihood of meeting criteria for PTSD. Higher household income was associated with reduced odds for a mental disorder.

In line with previous findings, respondents were more concerned about infecting others, than with being infected themselves (Shechter et al., 2020). Importantly, being concerned about passing COVID-19 on was a robust predictor of clinically significant disorders. Participants who had not had reliable access to PPE (approximately a third of participants) also had higher rates of clinically significant distress. These results emphasise the importance of providing adequate PPE to HSCWs throughout an infectious disease outbreak not only to protect their physical health but in order reduce the likelihood of mental distress. If HSCWs perceive themselves to be unsafe and vulnerable to contracting an infectious disease they may actively avoid loved ones in order to protect them (Billings, Ching, et al., 2020). Furthermore, it may be that others avoid socialising with or being in close proximity to HSCWs due to fear of infection, which may be experienced as rejection and stigmatisation. As a result, HSCWs are likely to have reduced social support from family and friends – a key protective factor for mental health - at a time when they have heightened levels of stress and distress.

Crucially, over 30% of participants reported that they could not tell their manager how they were feeling, and this was associated with the highest odds of distress across most models. In addition, being redeployed was a risk factor for both PTSD and anxiety. Recently, interventions aimed at improving healthcare team leaders’ awareness of mental distress among their staff, and most importantly their skills in engaging in supportive conversations have been implemented in some settings (Greenberg & Tracy, 2020). The findings from the current study support the need for these kinds of interventions, and they may be especially helpful for staff who are redeployed to new roles or teams and lack their usual support network. It is important to acknowledge, however, that there may be complicated reasons behind workers not feeling able to talk to managers which warrant exploration. Furthermore, HSCWs are likely to be best helped by a network of support, including managers, peers and professional mental health support (Billings, Greene, et al., 2020).

In the current study, we directly compared nurses and midwives, allied healthcare professionals, carers, clinical support staff. We found that nurses and midwifes were more likely to meet criteria for PTSD compared with carers and clinical support staff, and to meet criteria for at least one mental disorder compared with allied healthcare professionals and carers. Although previous studies also found nurses to be particularly distressed (Billings, Ching, et al., 2020; de Pablo et al., 2020; Preti et al., 2020), most examined differences between nurses and doctors, with little or no comparison with other HSCW groups. Notably, an examination of the levels of PTSD, anxiety and depression symptoms among the different groups in this study did not indicate that nurses and midwives were at highest risk; all had similarly high symptoms of PTSD, depression and anxiety symptoms compared with nurses. Furthermore, for the current study, most of the occupational groups (including pharmacists, doctors, cleaners, porters and administrators among others) were too small to be entered as separate comparison groups and were collapsed into one heterogeneous ‘other roles’ group. Thus, it is likely that among this ‘other group’ there were particular occupational groups that were also highly distressed, and possibly even more so than those groups in the current study. We recommend conducting further studies examining COVID-19 distress in HSCWs, especially with non-clinical staff working in health and social care settings, who have been typically neglected in research to date.

In contrast to some other studies, gender was not a significant predictor of distress and neither was identifying as Black, Asian or other ethnic minority. These findings should be treated with caution as the majority of participants identified as women (92%) and white (91%), and it may be that there was insufficient power to detect differences. We recommend that further studies oversample Black, Asian and other ethnic minority groups, especially as emerging evidence indicates they have been disproportionately affected by the COVID-19 pandemic in the UK (Rimmer, 2020). Additionally, unlike some studies, we did not find that working directly with patients who were suspected or confirmed to be infected was a significant predictor of distress. This may reflect the fact that during the period of data collection COVID-19 was relatively widespread, and so even HSCWs who were not directly working with COVID patients perceived that they were at risk.

There are some limitations to this study. Although the sample was reasonably large, it was a convenience sample recruited through social media, and therefore is vulnerable to self-selection bias. Thus, it is not a representative sample, and prevalence estimates for the whole HSCW population in the UK should not be derived from this study. Second, the questionnaires were self-report, rather than standardised clinician-administered diagnostic interviews. Third, we were not able to include other occupational groups in the comparisons due to the small numbers of participants in each group. Finally, although this is part of a longitudinal study for which data are currently being collected, the current paper reports only on the first wave of this study, therefore it is not possible to draw conclusions as to the direction of these relationships.

### Clinical implications

This study found evidence of high levels of distress among the participants. While these rates may be over-estimates, they are indicative of clinically significant need. For many, this acute distress will naturally resolve and should not be pathologised (Billings, Greene, et al., 2020; Lamb, Greenberg, Stevelink, & Wessely, 2020). Furthermore, there is a temptation to rush to put in place formal interventions, yet for many HSCWs this will not be necessary. Nevertheless, for some frontline HSCWs, their distress and other symptoms will be severe and have the potential to become chronic. These cases need to be detected early and treated promptly in order to protect the individual HSCWs and the overall functioning of the entire health and social care system during the pandemic. The findings that distress levels were relatively high across the different occupational groups and work settings emphasises the need to ensure services reach out to all these groups.

### Conclusions

This study highlights the importance of reliable access to PPE, not only for the physical safety of staff, but also to reduce their feelings that they are at risk of catching COVID, and crucially, the perception that they are a risk to others. It is important to examine the role that managers can play in reducing staff distress, especially for redeployed staff. Differentiating between those with temporary distress, and those who are on a trajectory for longer-term mental health problems, is a priority. Identifying risk factors for PTSD, depression and anxiety among HSCWs, and providing treatment for those who need it is critical given that subsequent waves of COVID-19 and other healthcare crises are inevitable.

## Data Availability

Researchers can contact the authors for details on how to request the data.

