## Supplementary material for "Predictors and rates of PTSD, depression and anxiety in UK frontline health and social care workers during COVID-19"

Supplementary material contains:

1. Supplementary Table 1 – Health and social care workers by role and setting

2. Group difference tests: PTSD, depression and anxiety symptoms by role, setting, and by other demographic variables

3. Correlation matrix

4. Supplementary Table 2- mean symptom scores for individuals included in analysis

**1. Supplementary table 1 – Health and social care workers by role and setting**

|  | Other Community Setting | Hospital | Nursing or care home | Other | **TOTAL** |
| --- | --- | --- | --- | --- | --- |
| Allied healthcare professional | 57 | 76 | 0 | 3 | **136** |
| Carer | 21 | 3 | 80 | 1 | **105** |
| Clinical support staff | 67 | 110 | 24 | 3 | **204** |
| Nurse or midwife | 131 | 321 | 43 | 9 | **504** |
| Other | 78 | 126 | 28 | 9 | **241** |
| **TOTAL** | **354** | **636** | **175** | **25** | **1190** |

**2a. ANOVAs - PTSD, depression and anxiety symptoms by job role**

| Outcome | F value | | P value | |
| --- | --- | --- | --- | --- |
| PTSD symptoms | | 3.64 | | 0.006 |
| Depression symptoms | | 2.80 | | 0.025 |
| Anxiety symptoms | | 2.77 | | 0.026 |
| *Results of Tukey Honest Significant Differences test (only showing significant differences)* | |  | |  |
| PTSD symptoms \| \| \| \|; Comparison \| difference \| lower \| upper \| P-value; Clinical support vs Allied healthcare \| 2.42 \| 0.59 \| 4.24 \| 0.003; Depression symptoms \| \| \| \|; Comparison \| difference \| lower \| upper \| P-value; Clinical support vs Allied healthcare \| 2.46 \| 0.32 \| 4.60 \| 0.014; Anxiety symptoms \| \| \| \|; Comparison \| difference \| lower \| upper \| P-value; Clinical support vs Allied healthcare \| 2.43 \| 0.40 \| 4.46 \| 0.009 | |  | |  |

**2b. ANOVAs - PTSD, depression and anxiety symptoms by work setting**

| Outcome | F value | | P value | |
| --- | --- | --- | --- | --- |
| PTSD symptoms | | 3.54 | | 0.014 |
| Depression symptoms | | 1.10 | | 0.346 |
| Anxiety symptoms | | 1.66 | | 0.174 |
| *Results of Tukey Honest Significant Differences test (only showing significant differences)* | |  | |  |
| PTSD symptoms \| \| \| \|; Comparison \| difference \| lower \| upper \| P-value; Nursing or care home vs other community setting \| 1.64 \| 0.21 \| 3.07 \| 0.017 | |  | |  |

**2c. t-tests – PTSD, depression and anxiety symptoms by demographic variables**

Independent-samples t-tests were conducted to see whether the binary demographic variables collected in the study (having children, relationship status, ethnicity and gender*) were associated with significantly different PTSD, depression or anxiety symptoms. None of the results were significant, with exception of having children and PTSD symptoms. The results of this independent samples t-test found that there was a significant difference in the ITQ scores for those with (M = 7.44, SD =5.53) and without children (M = 8.28, SD = 5.96), t(1088)=- 2.33, p = 0.020. This indicates that the individuals in the sample without children displayed higher PTSD symptoms.

*Note: For this analysis gender was transformed to a binary variable - the individuals who identified as “other” were excluded as this group was very small.

**3**. **Correlation matrix**

|  | Age | Income | Worried get COVID | Worried infect others | Stigma | PTSD symptoms | Depression symptoms |
| --- | --- | --- | --- | --- | --- | --- | --- |
| Age |  |  |  |  |  |  |  |
| Income | 0.11* |  |  |  |  |  |  |
| Worried get COVID | 0.035 | -0.08* | - |  |  |  |  |
| Worried infect others | -0.14*** | -0.09** | 0.54*** |  |  |  |  |
| Stigma | -0.16*** | -0.11* | 0.28*** | 0.28*** |  |  |  |
| PTSD symptoms | -0.09 | -0.14*** | 0.44*** | 0.43*** | 0.37*** |  |  |
| Depression symptoms | -0.11** | -0.17*** | 0.20*** | 0.30*** | 0.30*** | 0.60*** |  |
| Anxiety Symptoms | -0.13*** | -0.13*** | 0.27*** | 0.36*** | 0.27*** | 0.65*** | 0.80*** |

Note a: Spearman correlations

Note b: *p<0.05, **p<0.01, ***p<0.001

4. Supplementary Table 2- mean symptom scores for individuals included in analysis

691 (78.8%) of the individuals included in the depression model indicated that they work directly with COVID. This compares to 675 (79.5%) for anxiety and 744 for PTSD (78.9%).

|  | Direct work with COVID (Mean/SD) | |
| --- | --- | --- |
| Outcome | Yes | No |
| Depression | 10.2 (6.6) | 9.5 (6.6) |
| Anxiety | 8.6 (6.1) | 8.4 (6.2) |
| PTSD | 8.2 (5.9) | 7.3 (5.8) |
